# Kick the wheels: radiograph-negative ankle injuries in bicycle-spoke accidents

**DOI:** 10.1101/2022.07.26.22278082

**Authors:** Susanne JM Laumer, Lottie van Kooten, Dennis G Barten, Frits HM van Osch, Marion MWE Drees, Anita W Lekx

## Abstract

**Introduction:** Bicycle spoke injuries (BSIs) in children are notorious for the presence of Salter Harris type 1 (SH1) fractures. Most patients are therefore treated with cast immobilization. However, the actual prevalence of SH1 following a BSI is unknown. In this study, we aimed to describe a cohort with radiograph-negative BSIs and to identify possible clinical predictors for SH1 which might be useful for adequate risk assessment.

**Methods:** A retrospective cohort study was performed, including all children ≤12 years visiting our Emergency Department (ED) with a BSI. Patients without radiographic evidence of a fracture were classified as low or high level of suspicion of SH1. Multivariate logistic regression analysis was used to identify independent predictors of a high level of suspicion of SH1.

**Results:** In total, 323 patients were included. Ninety-three patients (29%) had a proven fracture; 230 patients were radiograph-negative at first presentation. Of these, 166 patients (72%) were treated with cast immobilization. At follow-up, 32 patients (13.9%) were classified as high level of suspicion of SH1. No clinical variables were found to be predictive for SH1. Local tenderness at the lateral malleolus was associated with a high level of suspicion of SH1, however, this was not statistically significant (OR 2.89, p-value 0.057).

**Conclusion:** Although BSIs with radiograph-negative ankle injuries are generally treated with cast immobilization, in this cohort only 13.9% had a high level of suspicion of SH1. Lateral malleolus tenderness was associated with a high level of suspicion of SH1 injury, but none of the clinical variables had a significant predictive value.

## Introduction

Bicycle spoke injuries (BSIs) are injuries of the lower leg resulting from entrapment of the leg between the frame and spokes of a rotating wheel. Because of the shearing injury mechanism, bicycle spoke accidents can lead to severe soft tissue lesions and fractures [1,2].

BSIs are frequently assessed in emergency departments (EDs) in cycling nations, such as the Netherlands, United States of America, India, China and Japan [2-5]. In the Netherlands, each year 3.000-5.200 children present to the emergency department (ED) with a BSI [6]. Young children (age 3-6 years) contribute to 2.200-4.200 of these visits [6]. Hence, BSIs are among the most common preventable injuries in young children. Preventive measures include the use of an appropriate child seat, spoke guards, pedals and foot straps [1].

Children with BSIs generally attend the ED because of pain caused by wounds, heel flap injuries, swelling or tenderness over the distal tibia, fibula or foot. Radiograph proven fractures are seen in about 30% of the cases [7-10]. The shearing injury mechanism often results in a forced inversion of the ankle [2,11,12]. Salter and Harris [13] reported that in children the epiphyseal plate is weaker than the surrounding tendons and ligaments. This results in a higher risk of epiphyseal fractures than injuries of the ligaments. Therefore, children with a BSI and local tenderness overlying the growth plate but who have no radiographic evidence of a fracture are often labelled as Salter Harris type 1 (SH1) fractures [12,13]. Even when there is a low level of suspicion, paediatric radiograph-negative ankle injuries are generally treated with cast immobilization for at least one week. However, the actual prevalence of SH1 following a BSI is unknown. A previous study [12] on children with non-specific ankle injuries, showed that the prevalence of SH1 was only 3% on magnetic resonance imaging (MRI), which is the golden standard to diagnose epiphyseal injures. Therefore, the current practice is probably associated with unnecessary cast immobilization and follow-up.

In this study, we aimed to describe a cohort with radiograph-negative BSIs and to identify possible clinical predictors for SH1 which might be useful for adequate risk assessment.

## Methods

### Study design and population

A retrospective cohort study was performed at the ED of a teaching hospital in the Netherlands. From January 2010 to December 2017 all children up to 12 years old visiting the ED with a BSI were included.

### Data collection

Data were collected from electronic patient charts (Chipsoft HiX, Amsterdam, the Netherlands), using a search engine that identified all records including the words ‘spoke’ or ‘spokes’. All records were subsequently screened for eligibility. Data extraction was independently performed by two researchers (SL and LK) to increase inter-rater reliability and several cases were reviewed twice to assess intra-rater variability. The following demographic and clinical data were registered: sex, age, referral type, affected side, ability bear weight, presence of lateral malleolus tenderness, presence of soft tissue injury (swelling, hematoma and/or abrasions), type of radiograph, diagnosis, initial treatment and follow-up. Patients with radiograph-negative ankle injuries at initial assessment in the ED were classified to have a low or high level of clinical suspicion of SH1 on the first follow-up appointment. The level of suspicion was based on initial assessment in the ED as well as the decision of the consulting physician to prolong cast immobilization on secondary assessment (typically seven days post-injury) in the outpatient setting. High clinical suspicion was defined as a high suspicion of SH1 at first assessment in the ED and prolongation of cast immobilization at outpatient follow-up. Low clinical suspicion was defined as low clinical suspicion at follow-up and the decision to not prolong immobilization. Due to the lack of golden standard imaging, which is MRI, high clinical suspicion of SH1 was used as a surrogate endpoint for an actual SH1 fracture. Follow-up duration was defined as the total number of days between ED presentation and the last outpatient visit. Furthermore, complications during follow-up such as infection, bleeding, cast problems or missed fractures were recorded.

### Data analysis

All data were analysed using SPSS, version 24. Descriptive data were analysed for all clinical variables using descriptive statistics. Continuous data were presented as mean ± standard deviation (SD), categorical data are presented as number and percentage. Patients with radiographic negative ankle injuries were divided into two groups: low or high level of clinical suspicion of SH1. Clinical variables were compared between these groups using the Chi-square test for categorical data and the Mann-Whitney U test for continuous data. Fisher’s Exact Test was used instead of the Chi-square test in case of small sample sizes. Clinical variables of which univariate analysis suggested a significant difference between the two groups were used as covariates in multivariate analysis. Multivariate logistic regression analysis was used to identify independent predictors of SH1.

## Results

A total of 323 patients were included. The mean age was 5 years and 56.7% was male. Ninety-three patients (29%) had a proven fracture; 230 patients were radiograph-negative at first presentation (Table 1). Of these, 166 patients (72%) were treated with cast immobilization, 38 (17%) with compression bandage, two (1%) required wound closure and 24 (10%) were not immobilized. At follow-up assessment 7 days post-injury, 32 patients (9.9%) of total study population and 13.9% of the radiograph negative cohort were eventually classified as high level of suspicion of SH1. Four patients (1.2%) who were not immobilized in the ED, received cast immobilization at subsequent follow-up (Figure 1).

**Table 1.** Patient characteristics.

|  | Total study population (n = 323) | Proven fracture (n = 93) | Radiograph-negative cohort (n = 230) |
| --- | --- | --- | --- |
| Age (years) | 5.2 ± 2.1 | 5.1 ± 1.7 | 5.3 ± 2.3 |
| Sex |  |  |  |
| Male | 183 (56.7) | 49 (52.7) | 134 (58.3) |
| Female | 140 (43.3) | 44 (47.3) | 96 (41.7) |
| ED referral |  |  |  |
| First line referral | 205 (63.5) | 61 (65.6) | 144 (62.6) |
| Self-referral | 118 (36.5) | 32 (34.4) | 86 (37.4) |
| Affected side |  |  |  |
| Left | 183 (56.7) | 47 (50.5) | 136 (59.1) |
| Right | 138 (42.7) | 44 (47.3) | 94 (40.9) |
| Both | 2 (0.6) | 2 (2.2) | 0 (0) |
| Ability to bear weight |  |  |  |
| Yes | 58 (18.0) | 10 (10.8) | 48 (20.9) |
| No | 169 (52.3) | 50 (53.8) | 119 (51.7) |
| Missing data | 96 | 33 | 63 |
| Soft tissue injury |  |  |  |
| Yes | 315 (97.5) | 89 (95.7) | 226 (98.3) |
| No | 5 (1.5) | 2 (2.2) | 3 (1.3) |
| Missing data | 3 | 2 | 1 |
| Lateral malleolus tenderness |  |  |  |
| Yes | 184 (57.0) | 59 (63.4) | 125 (54.3) |
| No | 89 (27.6) | 17 (18.3) | 72 (31.3) |
| Missing data | 50 | 17 | 33 |
| Treatment |  |  |  |
| Cast | 256 (79.3) | 90 (96.8) | 166 (72.2) |
| Bandage | 40 (12.4) | 2 (2.2) | 38 (16.5) |
| None | 24 (7.4) | 0 (0) | 24 (10.4) |
| Other | 3 (0.6) | 1 (1.1) | 2 (0.9) |
| Duration follow-up (days) | 17.8 ± 26.1 | 25.9 ± 19.2 | 13.9 ± 28.1 |
| Outpatient visits |  |  |  |
| 0 | 41 (12.7) | 5 (5.4) | 36 (15.7) |
| 1 | 139 (43.0) | 20 (21.5) | 119 (51.7) |
| >1 | 115 (35.6) | 61 (65.6) | 54 (23.5) |
| Missing data | 28 | 7 | 21 |
| High level of suspicion SH1 after outpatient visit | 32 (9.9) | - | 32 (13.9) |
| Complications | 38 (11.8) | 14 (15.1) | 24 (10.4) |
| Loss to follow-up | 67 (20.7) | 12 (12.9) | 55 (23.9) |

Continuous data are presented as mean ± SD; categorical data are presented as number (%). Missing values are indicated in italics.

**Figure 1.**
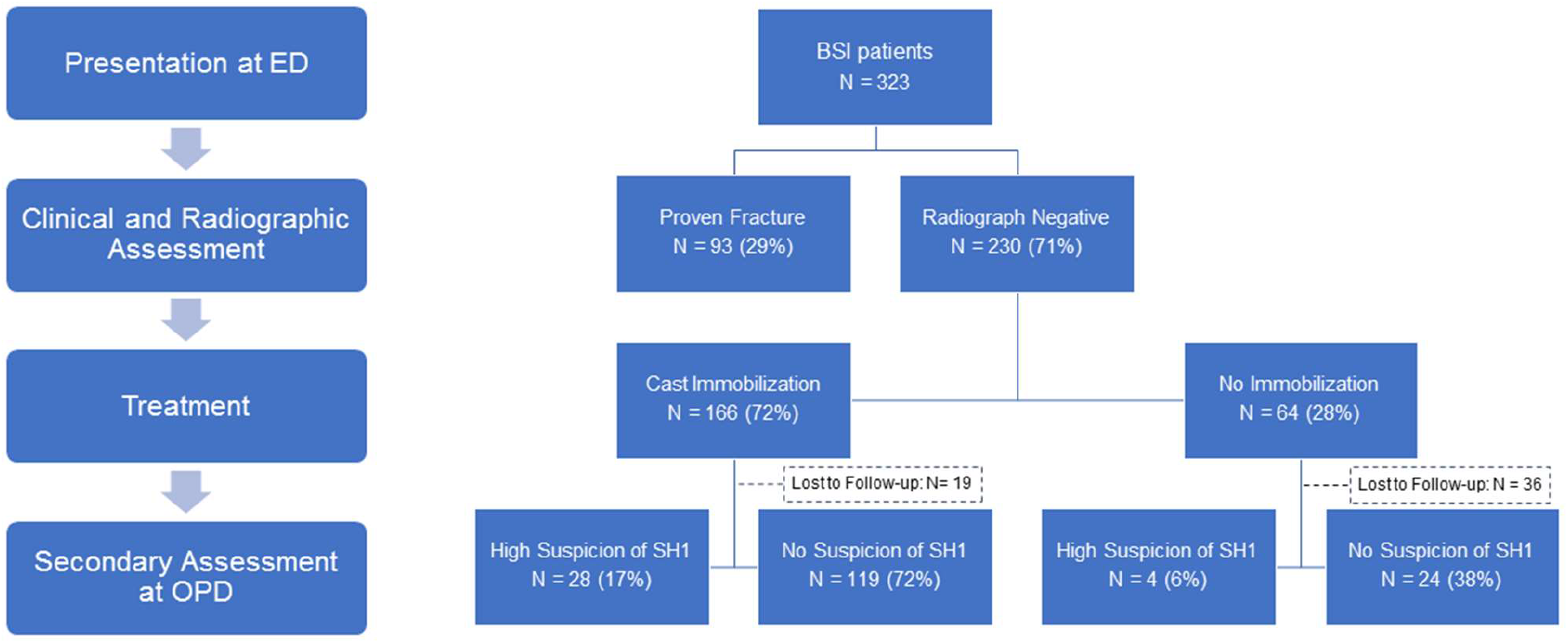
Flow-chart illustrating study design.

Median follow-up was fourteen days in the radiograph negative cohort. Within this group, 33 patients were lost to follow-up. Reported complications were: cast complaints (n= 4, 1.7%), impaired wound healing or wound infection (n=8, 3.5%) and extra outpatient clinic visits due to need for additional diagnostic tests (n=2, 0.9%) under treatment (for example: ongoing pain due to soft tissue injury or high suspicion of SH1) (n=8, 3.5%).

Clinical variables (Table 2) were compared between the groups “low and high suspicion of SH1” to identify predictors for SH1. None were found to be predictive. Lateral malleolus tenderness was more common in patients with high suspicion of SH1 (84.6% versus 65.6%), but statistical significance was not reached (OR 2.89, p-value 0.057). Affected side, presence of soft tissue injury and the ability to bear weight after BSI were not found to be associated with high suspicion of SH1. Follow-up duration was longer in the high level of suspicion of SH1 group (28.2 versus 10.7 days, p-value <0.001).

**Table 2.** Univariate Analysis of Independent Predictors of a SH1 fracture.

|  | High level of suspicion SH1<br>(n = 32) | Low level of suspicion SH1<br>(n = 143) |  |  |
| --- | --- | --- | --- | --- |
| | N (%) / mean ± SD | N (%) / mean ± SD | p-value | OR [95%CI] |
| Male | 18 (56.3) | 85 (59.4) | 0.740 | 0.877 [0.405-1.903] |
| Self-referral | 10 (31.3) | 50 (35.0) | 0.689 | 0.845 [0.371-1.925] |
| Left side affected | 21 (65.6) | 85 (59.4) | 0.518 | 0.768 [0.344-1.712] |
| Soft tissue injury | 32 (100) | 142 (99.3) | 0.817 | # |
| Able to bear weight after BSI (52) | 4 (22.2) [14] | 25 (23.8) [38] | 0.576 | 0.914 [0.276 – 3.031] |
| Lateral malleolus tenderness | 22 (84.6) | 80 (65.6) | 0.057 | 2.888 [0.934-8.029] |
| Follow-up duration (days) | 28.2 ± 61.9 | 10.7 ± 8.8 | <0.001<br>(781, -5.575)* | 0.939 [0.902-0.977] |
| Outpatient visit (>1) | 27 (84.4) | 27 (19.3) | <0.001 | 22.600 [7.968 – 64.101] |

Number between brackets in first column is number of missing values; in second and third column missing values are between []. \*U and Z-scores are presented between brackets resp. (fourth column). OR = Odds ratio. # not calculated since approximately 100% of included patients presented with soft tissue injuries in both categories.

A multivariate logistic regression analysis was performed to adjust for the influence of possible confounders regarding the association of lateral malleolus tenderness with high suspicion of SH1. Multivariate analysis for all possible confounders yielded a significant difference, but the confidence interval became very wide due to missing values which resulted in unreliable results. Therefore, we adjusted only for age and gender. The multivariate adjusted OR was 2.81 (CI 0.868 – 9.102).

## Discussion

In this study to assess radiograph-negative ankle injuries in BSI patients, 71% of BSI patients were radiograph-negative and 29% had a radiologic proven fracture of the lower extremity at initial assessment. Of all radiograph-negative BSIs, 72% received cast immobilization. Only 14% was eventually classified as a high clinical suspicion of SH1 at follow-up. Even though lateral malleolus tenderness was associated with a high suspicion of SH1, statistical significance was not reached.

In a similar study from the Netherlands [7], the researchers also aimed to identify clinical signs that could predict the presence of a fracture. However, patients without evidence of a fracture were compared to patients with a proven *and* a possible fracture (including SH1). Our study particularly focused on the radiograph-negative cohort as there is no doubt about the treatment of patients with radiograph proven fractures. In both studies, no significant clinical predictors for proven fractures and SH1 were found.

A radiograph-proven fracture was found in 29% of all BSI patients, which is consistent with previous research on BSIs [7-10]. Fourteen percent of the patients with radiograph-negative ankle injuries had a high suspicion of SH1. This is relatively high compared to the findings of a study on radiograph-negative lateral ankle injuries by Boutis et al., where the incidence of SH1 was confirmed by MRI in only 3% of patients [12]. Possibly, the relatively high proportion of ankle injuries with high suspicion of SH1 in our cohort might be explained by the higher rate of soft tissue injuries that are associated with the shearing injury mechanism of BSIs. The MRI confirmation study assessed all lateral ankle injuries and not exclusively assessed BSIs.

Complications were found in 10% of all patients; only 1.7 percent seemed to be directly related to cast immobilization. Moreover, all complications were considered to be mild. More severe consequences of cast immobilization, such as thermal injury, pressure sores, infection and neurovascular injury [14] are rarely reported, but these risk should be weighed up against the numerous such as pain management, promotion of wound healing and the prevention of pes equines [10]. Based on the complication rate alone, cast immobilization for BSIs should not be discouraged.

The mean follow-up in our study was shorter compared to previous studies; 13.9 days versus 17.4 days [10]. Therefore, long-term consequences of BSIs were not assessed. Other studies showed that the incidence of persistent medical complaints after a BSI, both physical and psychological, is relatively high. For example, persistent psychological problems after a BSI are reported in 50% of children [9]. This highlights the importance of prevention of these injuries.

### Strengths and limitations

A key strength of the study is the large study population of 323 patients, with a subgroup analysis of 230 radiograph-negative patients.

However, this study also encompasses numerous limitations that need to be acknowledged. First, the retrospective study design is prone to information bias. This single-centre study concerns a retrospective analysis of written and digital patients records from which several limitations arise, such as under-registration or misclassification.

Second, there is a certain degree of loss to follow-up (22.9% in radiograph-negative cohort), in which the dropout rate differs between the “cast immobilization” (11.4%) and “no immobilization” (56.3%) group. Reasonably, patients without cast immobilization are more likely to be lost to follow-up due to the fact that they do not need to have the cast removed at the outpatient department. Furthermore, the decision of the emergency physician to not immobilize the patient suggests that they had only minor complaints at initial presentation at the ED. Still, we could not allocate these patients to the low suspicion of SH1 group (and thereby lowering the loss to follow-up rate) because the patients might have had an outpatient appointment in a different hospital.

As previously stated, no clinical variables were found to significantly predict SH1 in BSI. Nonetheless, lateral malleolus tenderness was associated with a high suspicion of SH1, with a p-value approaching 0.05 and an OR of almost 3. Given the relatively small cohort of 32 patients with high suspicion of SH1 this suggests a strong correlation. Probably, a larger study population and a prospective study design with a more detailed and formalized description of the physical exam would have resulted in statistically significance.

Nevertheless, it is important to pay careful attention to the lateral malleolus during physical examination.

Finally, the final diagnosis of SH1 was not verified by the gold standard, which is MRI. Therefore, the actual prevalence of SH1 is probably lower, but unknown. This subjective assessment possibly resulted in a more heterogeneous group with “high suspicion of SH1” complicating the identification of clinical predictors of SH1.

## Conclusion

Although BSIs with radiograph-negative ankle injuries are generally treated with cast immobilization, only 14% had a high level of suspicion of SH1. In this study, no clinical variables could significantly predict a high level of suspicion of SH1. However, lateral malleolus tenderness was associated with a high clinical suspicion of SH1 injury. Future studies are warranted to further improve risk assessment and treatment of BSIs.

## Supporting information

STROBE checklist cohort studies

ICMJE disclosure form

## Data Availability

All data produced in the present study are available upon reasonable request to the authors.

## Notes

### Competing Interest Statement

The authors have declared no competing interest.

### Funding Statement

This study did not receive any funding.

### Author Declarations

The Institutional Review Board of VieCuri Medical Centre waived ethical approval for this work.

